# Utilization of the Community Engagement Studio model to facilitate participatory design of a multicenter randomized controlled trial of the efficacy of scoliosis specific exercise rehabilitation

**DOI:** 10.64898/2026.01.06.26343535

**Authors:** Sanja Schreiber, Patricia J Piechowski, Amber Basnaw, Knoll Larkin, Dominique L Kinnett-Hopkins, Anna L Kratz, Emily C Somers

## Abstract

**Introduction:** Engaging individuals with lived experience enhances the rigor and real-world relevance of clinical research. In adolescent idiopathic scoliosis (AIS), where non-surgical evidence remains limited, involving patients and caregivers informs design, recruitment, and retention for future trials.

**Methods:** Using the Community Engagement (CE) Studio model, we conducted a 1.5-hour virtual session with children aged 10-16 years with AIS and their caregivers residing in North America. Participants (no spinal surgery; with/without brace) were recruited through engagement databases, advocacy organizations, and social media. A trained facilitator led structured discussions. Responses were thematically synthesized, including summaries related to recruitment, retention, study protocol, meaningful outcomes and treatment success. Participants received compensation and a follow-up Qualtrics survey. The CE Studio, based on the Vanderbilt model is advisory, and IRB- exempt.

**Results:** Of 81 respondents, 31 were eligible and 18 participated (nine girls, nine caregivers) representing seven ethnicities across nine U.S. states. All participants valued the proposed multicenter RCT on scoliosis exercise rehabilitation and expressed willingness to enroll; 78% would accept randomization to either the active or 6-month waitlist control arm. Reported barriers included limited access to physiotherapy (43%), physician skepticism (57%), and bracing preference (43%). The most bothersome symptoms were pain (50%) and brace discomfort (17%). Prioritized outcomes included preventing curve progression, avoiding surgery, pain reduction, and improved appearance.

**Conclusions:** Participants expressed strong willingness to enroll, emphasizing pain, progression, and access to care as key barriers. Improved communication with providers about non-surgical options was viewed as essential to support shared decision-making.

## Background

Adolescent idiopathic scoliosis (AIS) is a complex, progressive musculoskeletal condition that affects up to 5% of adolescents globally[1] and has a strong female predominance[2]. Profound implications for physical function, psychological well-being, and long-term health outcomes are associated with AIS. Despite its prevalence and clinical seriousness, scoliosis remains under-recognized as a public health and research priority and is inconsistently managed within healthcare systems.

In many regions, school-based screening programs have been discontinued or never implemented[3], and pediatric screening practices vary significantly [4]. As a result, an estimated 30% of children are referred to specialist care only after significant curvature progression, reducing the effectiveness and access related to non-invasive management options.[5,6]

Once within the clinical care pathway, adolescents with scoliosis are typically subject to frequent radiographic monitoring and presented with limited treatment options. These usually include rigid bracing for moderate curves or spinal fusion surgery for more severe cases.[7] Bracing is often associated with discomfort,[8] social stigma,[9] and adherence challenges, while surgery entails substantial risks and long-term consequences for spinal mobility and quality of life[10]. The period between diagnosis and initiating a treatment plan is frequently marked by emotional distress, as families navigate uncertainty, extended wait times, and fragmented access to care.[11]

There is increasing global interest in scoliosis-specific exercise rehabilitation (SSER) as a conservative, patient-centered complementary option to conventional approaches for mild or moderate AIS to reduce risk of progression to the surgical range and improve patient-reported outcomes.[12] However, despite promising early findings, much of the existing literature on exercise-based management remains limited by methodological weaknesses, including small sample sizes, lack of standardized protocols, inconsistent outcome measures, and inadequate follow-up.[13] This has contributed to variability in clinical uptake and a lack of integration into mainstream care. Nonetheless, the increasing demand for conservative options reflects a notable trend: patients and caregivers are frustrated with “watchful waiting” approaches and actively seek proactive, evidence-informed, and less invasive approaches to scoliosis care that prioritize quality of life and long-term function, and that align more closely with their values and preferences.[11,14,15]

Equally concerning are the variations in access and outcomes shaped by social elements. In both publicly funded and privatized health systems, factors such as race, insurance status[16], and geographic location influence the timing of diagnosis[17], treatment eligibility, and overall experience of care. Adolescents from under-resourced communities often present with more advanced curves at diagnosis, face longer surgical wait times, and are less likely to receive early intervention or consistent follow-up.[17] These differences highlight the need for more egalitarian, inclusive research frameworks that integrate varied perspectives and lived experiences into study design and implementation.

Collaborative partnerships between researchers and those with lived experience, including patients, families, and community partners ensures that research is not only scientifically rigorous but also grounded in the reality of those it seeks to serve. In the context of AIS, where current care pathways can be disempowering and reactive, engaging the community offers an opportunity to co-design interventions that are more acceptable, feasible, and aligned with patients’ priorities.

The Community Engagement (CE) Studio model provides a structured framework to facilitate this process. Developed by the Meharry-Vanderbilt Community Engaged Research Core (CERC) [18], the CE Studio enables researchers to gather timely, targeted input from *community* experts at various stages of a project, helping shape more effective support structures, behavioral strategies, and flexible intervention designs that better align with real-world experiences and constraints.

We applied the CE Studio model to inform the participatory design of a multicenter randomized controlled trial (RCT) evaluating the efficacy of scoliosis-specific exercise rehabilitation for adolescents with AIS. This manuscript outlines the rationale for this engagement, describes the CE Studio process, and shares insights on barriers, facilitators, and strategies for recruitment, retention, outcome selection, and defining success gained from our initial session. By embedding perspectives of those with lived experiences from the outset and throughout all steps of the trial, we aim to enhance its relevance, inclusivity, and translational potential, contributing to more responsive, person-centered scoliosis care.

## Methods

### CE Studio design and framework

The CE Studio model, which offers a structured framework to support participatory design, was conducted with children living with AIS and their caregivers. The CE Studio was implemented with trained facilitators, following the model described by the Meharry Vanderbilt Community Engaged Research Core [18]. Unlike traditional focus groups, which primarily aim to collect qualitative research data, CE Studios function as consultative, bi-directional sessions in which community members offer experiential insights to guide the development of research protocols.

### CE Studio rationale

The CE Studio model is designed to facilitate meaningful engagement between researchers and community stakeholders, including patients, families, and caregivers, ensuring that study designs are scientifically rigorous, relevant, and feasible within real-world contexts. This is particularly critical in AIS, where treatment pathways often fail to incorporate the lived experiences of affected individuals and where non-surgical rehabilitation options remain underdeveloped. The CE Studio framework supports input on aspects such as recruitment strategies, intervention feasibility, adherence challenges, and outcome measures, while also fulfilling funding agencies’ expectations for inclusive, patient-centered research design.

### CE Studio preparation and facilitation

The CE Studio was conducted by a facilitation team trained in this Vanderbilt model, and included the following core components:

1. Initial consultation between the research team and CE Studio facilitators to refine project goals.
2. Recruitment of a tailored panel of Community Experts aligned with the AIS patient population.
3. Pre-meeting coaching and orientation for researchers to prepare presentations and questions.
4. Pre-testing and refinement of questions with a group of six adolescents, with and without scoliosis, who were not a part of the Community Expert panel.
5. Virtual orientation session for Community Experts to familiarize them with the CE Studio process.
6. A 90-minute, facilitator-led virtual session during which the research team presented the study overview and posed structured questions.
7. Real-time documentation of participant feedback, including individual suggestions and thematic input.
8. Compensation for Community Experts to recognize their time and expertise.
9. A post-session survey, delivered via Qualtrics one week after the Studio, to gather additional reflections and validate insights.

### CE Studio community experts/participants

Eligible participants for the CE Studio were children aged 10–16 years living with AIS (with or without bracing), who had not undergone spinal surgery, and/or their primary caregivers, and residing in the US or Canada. This inclusive approach ensured a breadth of perspectives reflective of the study’s target population. Community Experts were recruited through several pathways: prior engagement databases, scoliosis-focused community organizations (*e.g.*, Higgy Bears and Scolios-Us), and targeted outreach via social media groups. This multi-faceted recruitment strategy aimed to maximize diversity in experiences, socioeconomic background, and treatment exposure.

### CE Studio session implementation

The CE Studio was conducted as a single 1.5-hour virtual Zoom session [19]. After a brief introduction, the research team presented an overview of the proposed trial, including the intervention, study goals, and key design components. The facilitator then guided a structured discussion using pre-developed prompts to elicit participant feedback on recruitment, intervention delivery, outcome prioritization, and anticipated barriers. All contributions were documented using structured note-taking templates to capture emerging themes and specific recommendations, and qualitative analysis software was not used. Participants were compensated with a consulting fee in recognition of their contributions.

### CE Studio content

To guide the structured discussion during the CE Studio, the facilitation team developed a series of open-ended, participant-centered questions designed to elicit meaningful feedback on study design, feasibility, and perceived value from the perspective of individuals living with AIS and their caregivers. As part of the development process, the set of proposed questions underwent review and refinement in an informal focus group of six adolescents (who were not part of the upcoming Community Expert panel), with and without AIS, to ensure clarity of wording and comprehension in that age group.

The discussion questions were tailored to explore motivations for study participation, treatment expectations, and personal experiences with scoliosis. The discussion aimed to surface both individual insights and common themes that could inform recruitment strategies, outcome prioritization, and intervention acceptability. CE Studio participants were asked to reflect on the prompts presented in Table 1, organized by thematic domain.

**Table 1.** CE Studio Discussion Questions.

| Thematic Domain | Discussion Questions |
| --- | --- |
| **Study Enrollment and Motivation** | • If you were invited to participate in this study, knowing that you might start the exercises now or in 6 months, why would you or wouldn't you choose to participate?<br>• What would make you want to or not want to be in the study? |
| **Control Group Considerations** | • Would you be willing to hold off from other scoliosis exercise treatments during the 6-month wait time (for the control group)? |
| **Lived Experience with Scoliosis** | • Does your scoliosis affect your day-to-day life, and if it does, how so?<br>• When thinking about your scoliosis, what, if anything, is bothering you the most about it? |
| **Treatment Goals and Success Criteria** | • What would make the treatment successful to you?<br>• How would you know that the treatment is successful?<br>• What are your goals for this treatment? |

These questions were asked in a conversational, non-directive manner to foster openness and encourage diverse perspectives. The facilitator ensured equitable participation and created space for both children and caregivers to share their views, either individually or collaboratively. This content formed the core of the CE Studio dialogue and shaped subsequent refinements to the trial protocol.

### CE Studio post-session feedback

To supplement insights gathered during the session, a follow-up survey was distributed to participants one week later using Qualtrics. The survey included both open-ended and structured items, allowing participants to elaborate on their feedback or offer reflections they may not have shared during the live session.

### Ethical considerations

As a consultative model, CE Studios are considered advisory and not research. Thus, they do not require IRB approval. Participants were informed of the nature and purpose of the session, and no identifiable or sensitive data were collected or analyzed. Participation was voluntary, and all community experts received consulting honoraria for their time and insights.

## Results

Following a two-week recruitment period through multiple community outlets, a total of 81 individuals responded to the invitation. Of these, 31 met the eligibility criteria, residing across 16 U.S. states and one Canadian province. A final cohort of 18 Community Experts participated in the CE Studio, comprising nine adolescent girls with AIS (aged 12 to 16 years) and their caregivers. Participants self-identified across seven ethnicities and were geographically distributed across nine U.S. states, contributing multifaceted perspectives. Structured notes revealed four distinct themes (Table 2).

**Table 2.**
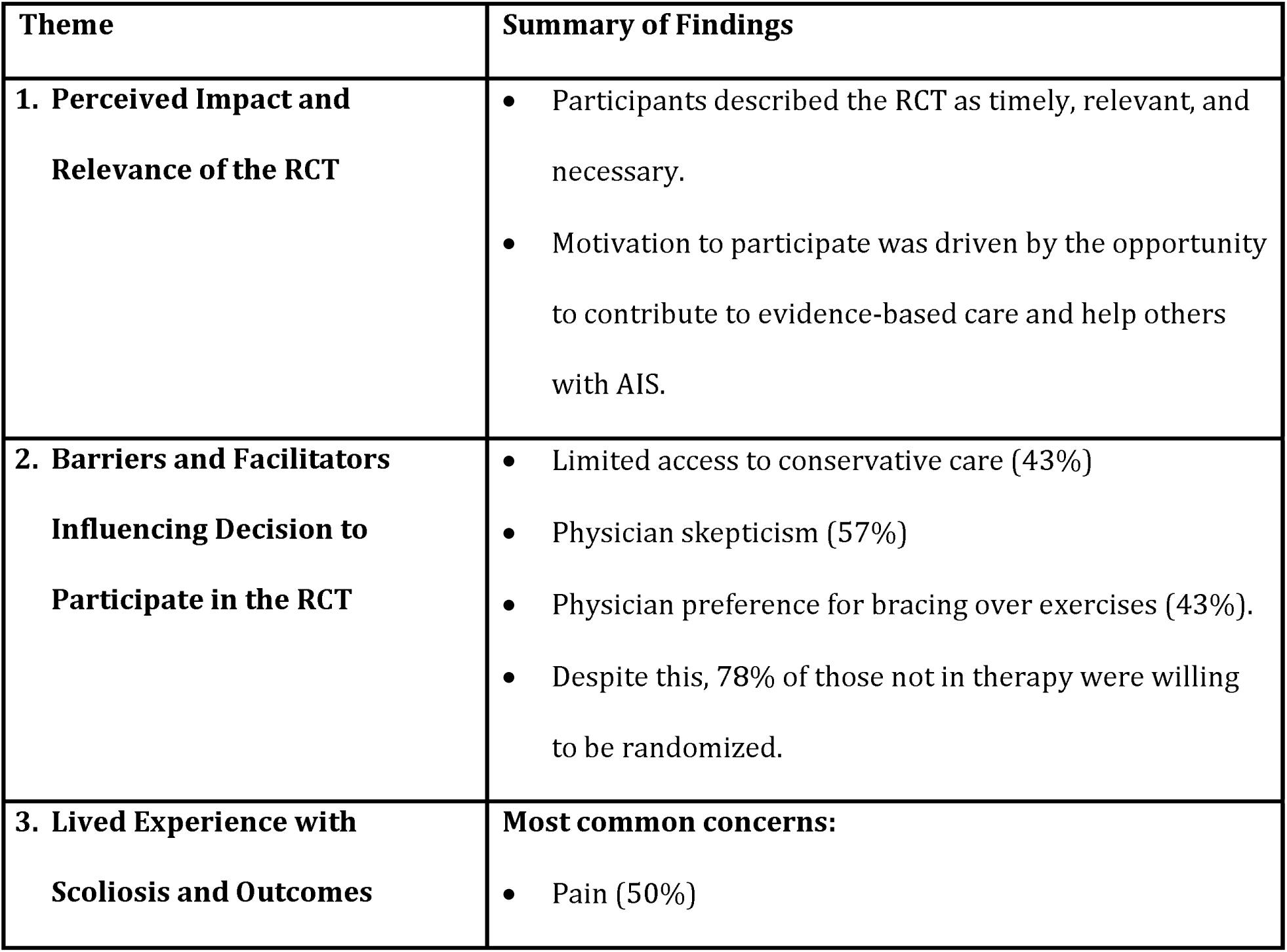
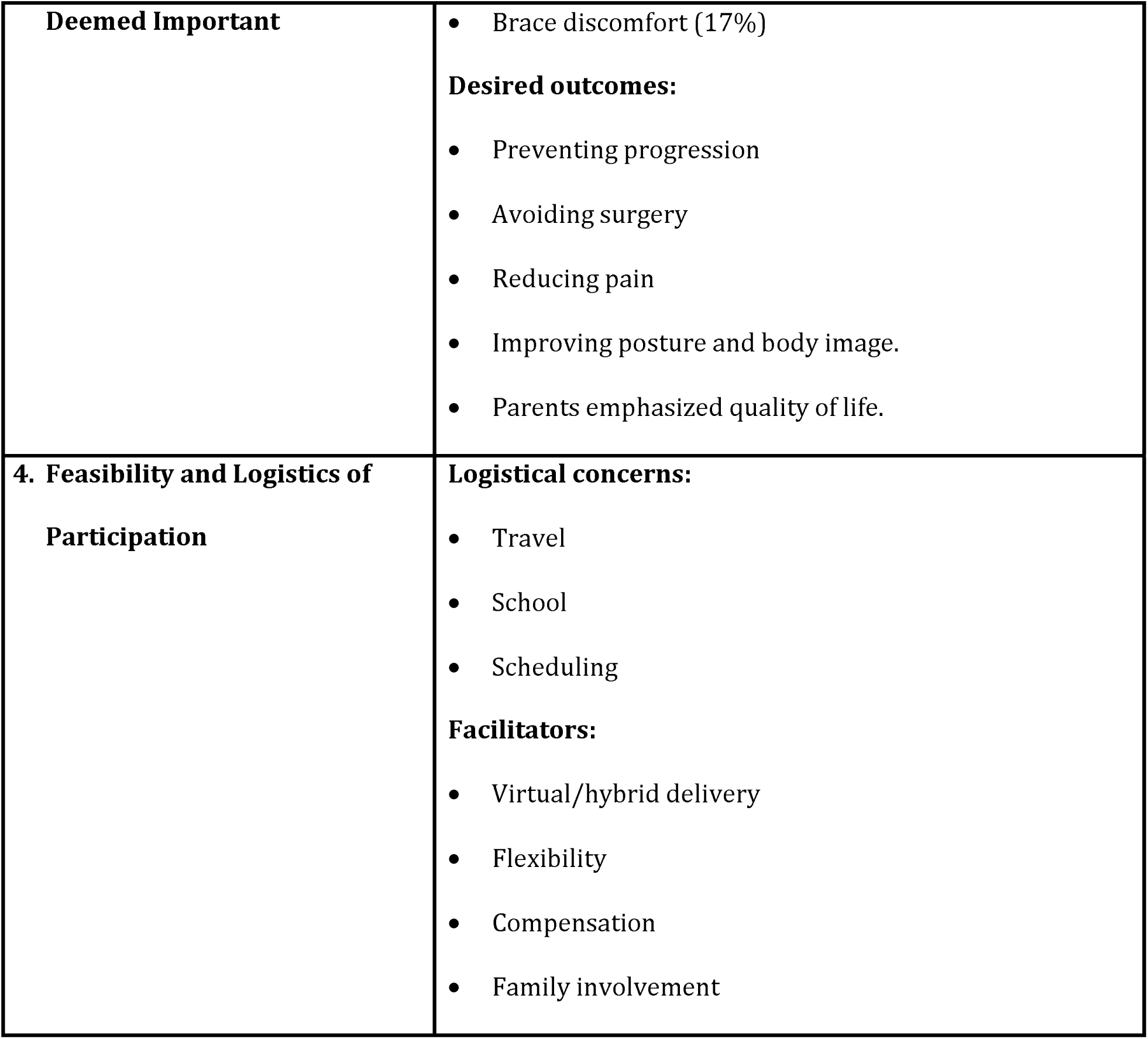
Themes and Summary of Findings.

### Theme 1: Perceived Impact and Relevance of the RCT

All participants expressed strong interest in the proposed RCT and described it as timely, relevant, and valuable. The perceived importance of the research and the opportunity to contribute to improved treatment options for others were commonly cited motivations for participation.

### Theme 2: Barriers and Facilitators Influencing Decision to Participate in the RCT

Among those not currently engaged in scoliosis-specific physical therapy or rehabilitation, several *barriers* were identified:

- 43% reported limited access to conservative care in their area
- 57% cited skepticism or lack of recommendation from their physicians
- 43% stated that their physicians recommended bracing as the only viable option compared to physical therapy/rehabilitation exercises.

Despite these challenges, 78% of respondents not in current physical therapy indicated they would be willing to be randomized, either to begin the exercise program immediately (at the RCT baseline) or to wait six months as part of a control arm. Willingness to delay treatment was influenced by trust in the study design, belief in the program’s potential benefit, and recognition of the importance of evidence-based approaches.

### Theme 3: Lived Experience with Scoliosis and Outcomes Deemed Important

Pain emerged as the most frequently reported symptom impacting daily life, cited by 50% of the adolescent participants. An additional 17% mentioned discomfort and lifestyle limitations associated with bracing. Adolescents described physical restrictions, self-consciousness, and reduced participation in physical or social activities as secondary challenges.

Participants consistently identified the following as key desired outcomes from the intervention:

- Prevention of curve progression
- Avoidance of spinal fusion surgery
- Reduction in pain
- Improvements in posture and body image

Caregivers emphasized broader quality-of-life concerns, including emotional well-being, social participation, and treatment burden. Both adolescents and parents articulated a preference for proactive, non-invasive interventions that could delay or replace the need for bracing and/or surgery.

### Theme 4: Feasibility and Logistics of Participation

Participants raised logistical considerations related to travel, scheduling, and school conflicts but viewed them as manageable, particularly if the program could be offered in a hybrid or fully virtual format. Most participants expressed confidence in their ability to adhere to the intervention schedule if provided with structured support and flexibility. Compensation, clear communication, and continued family involvement were noted as facilitators for long-term engagement.

## Discussion

The CE Studio model offers a practical and impactful approach to incorporating lived experience into scoliosis research. By utilizing the CE Studio early in the clinical trial planning process, we obtained structured input from patients and caregivers, helping us revise and refine our protocol, including defining study outcomes, accordingly. This approach has the potential to shift the paradigm of AIS research toward more patient-centered, collaborative, and responsive models of care, addressing long-standing gaps in conservative scoliosis management and supporting more effective shared decision-making in clinical practice.

### Strengths

This study represents a novel application of the CE Studio model in the context of AIS, a condition where conservative care remains underutilized and under-researched. The structured engagement of adolescents with AIS and their caregivers at the pre-trial design stage allowed for the integration of lived experience into the development of a multicenter RCT protocol on scoliosis-specific exercise rehabilitation. This is particularly important in a field where treatment pathways are often prescribed without substantial input from patients and their families.

By using the CE Studio model, we were able to explore key feasibility questions around study enrollment, acceptability of randomization (including delayed intervention), and willingness to forego alternative therapies during the control period. Importantly, the CE Studio highlighted real-world barriers to accessing scoliosis-specific exercise rehabilitation, including the scarcity of trained providers, physician skepticism toward non-bracing conservative approaches, and beliefs about bracing superiority along with lack of appreciation for the role of exercise rehabilitation in conjunction with bracing - factors that are often overlooked in conventional study planning.

The inclusion of participants from diverse geographic regions and multiple ethnicities enhanced the relevance of the findings across varied healthcare contexts. Furthermore, the CE Studio provided valuable insights into patient-centered outcomes, including pain, self-image, and quality of life, which are often overlooked in clinical management and research that focus primarily on the curve severity outcome (Cobb angle).

Finally, an important strength of the CE Studio approach is that it established the foundation for ongoing, intentional follow-ups with experts. This ensures that patient and caregiver perspectives will continuously guide our research process beyond the planning phase, through trial implementation, interpretation of findings, and ultimately the dissemination and knowledge translation stages.

### Limitations

Despite these strengths, the CE Studio had several limitations. Participants were self-selected through community outreach and advocacy networks, which may have introduced sampling bias. Those who chose to participate may have been more motivated to seek non-surgical options or more engaged with scoliosis advocacy, potentially limiting generalizability to all AIS patients and families, particularly those who are less informed or connected to scoliosis-specific resources.

Additionally, while the CE Studio facilitated meaningful discussion, it was not designed as a formal qualitative study. Transcripts were not coded using qualitative analysis software, and thematic saturation was not assessed. As such, findings should be interpreted as formative, guiding trial design rather than producing generalizable conclusions.

Finally, the session captured input at a single time point, prior to trial launch. Ongoing planned engagement with community experts throughout the study lifecycle, including during recruitment, intervention delivery, and dissemination, is planned to sustain patient-centeredness and ensure continued alignment with participant priorities.

## Conclusion

The CE Studio process revealed strong support among adolescents with AIS and their caregivers for participating in an RCT of scoliosis-specific exercise rehabilitation, including willingness to be randomized to either the intervention or control arm. The most frequently cited concerns were pain and curve progression, with a consistent emphasis on avoiding surgery. Participants identified significant barriers to accessing conservative therapy, including limited provider availability and insufficient guidance from healthcare professionals on non-operative management options. These findings highlight a pressing need to improve communication, expand access to evidence-based conservative care, and integrate patient perspectives into clinical decision-making for AIS.

## Data Availability

NA

## Acknowledgments

The authors wish to thank the entire Community Engagement CE Studio Team at the Michigan Institute for Clinical & Health Research (MICHR), which comprises members of the Community Engagement, Patient Partners, and Participant Recruitment & Retention Programs. A big thank you goes out to the children, who are living with adolescent idiopathic scoliosis, and their families who participated in the CE Studio. Their amazing input was thoughtful, insightful, and impactful.

